# Patient personas exemplifying behavioural barriers and enablers to patient education across the hypertension journey: A qualitative framework analysis

**DOI:** 10.64898/2025.12.10.25341407

**Authors:** Kaylee Slater, Eleanor Clapham, Kim Beesley, Carissa Bonner, Elizabeth Halcomb, Ben Kostyrka, Lisa Kouladjian O’Donnell, Liliana Laranjo, Florence Lopez, Mitchell Sarkies, Gautam Satheesh, Mouna Sawan, Catherine Stephen, John Stevens, Ritu Trivedi, Aletta E Schutte, Niamh Chapman

**Author notes:** **Corresponding author:** Dr Niamh Chapman, School of Health Sciences, Faculty of Health and Medicine, The University of Sydney, Australia. Joint-first authors (Kaylee Slater, Eleanor Clapham).

## Abstract

**Background.:** Only one in five people with hypertension have their blood pressure (BP) controlled (<140/90 mmHg). Patient education improves BP control, yet there is limited guidance to adapt to individuals’ evolving needs across the hypertension management journey. This study aimed to identify key behavioural barriers and enablers to patient education throughout the hypertension management journey, which were synthesised into patient personas to support translation and practical application.

**Methods.:** Qualitative interviews with adults (≥18) who self-monitor BP, and primary care providers involved in hypertension care. Interviews explored the experience of accessing and delivering patient education for BP measurement and management. Using framework analysis, patient personas were developed in three steps: 1) thematic analysis of patient interviews to identify key barriers and enablers to hypertension patient education and mapping the identified barriers and enablers to key time-points in the hypertension journey (diagnosis, treatment initiation, long-term management), 2) clustering behavioural factors via the capability, opportunity, motivation-behaviour model, which were then synthesised as preliminary patient personas and refined using practitioner interviews and, 3) validation by consumer consultation feedback sessions.

**Results.:** Patients (n=27) and practitioners (n=12; n=4 general practitioners, n=4 nurses, and n=4 pharmacists) were aged 18-60 years (52% of patients and 100% of practitioners fell within this age range). Several behavioural barriers to patient education for hypertension management included patient overwhelm, inconsistent guidance provided by health professionals, and perceived patient disengagement, while a strong desire to self-manage was a consistent enabler. Six clusters of behavioural barriers and enablers were synthesised as patient personas, capturing the factors that shape education needs and experiences across the hypertension management journey.

**Conclusions.:** The patient personas highlight opportunities for tailored patient education strategies through the development of patient personas.

**Practice implications.:** The patient personas provide a practical tool for designing person-centred interventions in primary care.

## INTRODUCTION

High blood pressure (BP) is the global leading risk factor for mortality (1). Optimal BP control (<140/90mmHg) reduces the risk of associated morbidity and mortality; (2) yet only 20% of the 1.6 billion people with hypertension globally have their BP controlled (1).

Achieving BP control requires patients to undertake management behaviours such as lifelong medication adherence, lifestyle change (including diet and physical activity), long-term self-monitoring (home BP measurement) and navigation of multiple health services (3). Patients who receive education to support hypertension management achieve greater BP control (4) and have improved self-efficacy (5). This is because patient education empowers patients with the knowledge needed to engage in behaviours and decision-making to effectively manage their own health (6–10). As such, patient education is recognised within Roadmaps and Calls to Action globally as being integral to achieving BP control (6–9).

Hypertension is the most managed condition in primary care (11), which is therefore a critical setting for the delivery of patient education. In primary care settings, patients achieve greatest BP control outcomes when health education is delivered from healthcare practitioners (12, 13) through a collaborative, team-based approach involving general practitioners (GPs), nurses and pharmacists (14). However, healthcare practitioners receive minimal training to effectively deliver patient education that addresses patient needs (15). Further, opportunities for patient education in clinical settings are hindered by resourcing, time and funding constraints (16, 17). This results in patients looking elsewhere for this support and existing resources available to support patient education are unstandardized, do not meet patient needs and are not co-developed with people with lived experience of hypertension management (18).

Patients with hypertension have evolving informational needs across their care journey from diagnosis to long-term management (19), but these needs are not always well understood or addressed in practice (20). While primary care practitioners are well-positioned to provide tailored education, there is limited guidance on how to align education with patients’ changing needs. Inadequate patient education can contribute to poor BP control (4) and with increasing workforce pressures in primary care, understanding how to tailor education to changing patient behaviours and contexts is more critical than ever. Patient personas developed through qualitative research and co-design offer a creative way to capture and communicate diverse experiences, guide education delivery and promote patient-centred care by bringing patient perspectives to life in a tangible and relatable way (20, 21). Therefore, the aim of this study was twofold; 1) identify key behavioural barriers and enablers to patient education throughout the hypertension management journey, and 2) synthesise these factors into patient personas to support translation and practical application.

## METHODS

### Study design

This qualitative study included semi-structured interviews with patients and primary healthcare practitioners to explore patient education experiences in hypertension management. The University of Tasmania Human Research Ethics Committee (H0028867) and the University of Sydney Human Research Ethics Committee (2024/HE000528) gave ethical approval for this work. This study was reported as per the Consolidated Criteria for Reporting Qualitative Research (COREQ, Supplementary File 1) (22).

### Participant eligibility and recruitment

#### Patients

Adults aged ≥18 years, who measured BP at home in the past 12-months were recruited from a survey of home BP management (23) to participate in semi-structured interviews using an information power purposive sampling approach (24).

Participants were selected to have a diverse set of characteristics including location, gender, education level, and health literacy measured using a validated 3-item instrument (25). Participants had the opportunity to enter a raffle prize draw at the time of recruitment.

#### Practitioners

Primary healthcare practitioners (GPs, practice nurses/nurse practitioners and community pharmacists) who provided care to adults with hypertension in Australia were recruited to participate in semi-structured interviews using an information power purposive sampling approach (24). Practitioners were recruited through advertisements on Primary Health Network newsletters, email correspondence from supporting organisations to which they had previously subscribed, social media advertisements and convenience sampling. Practitioners were reimbursed to compensate for the time taken to participate.

### Data collection

#### Participant characteristics

Demographic information was collected from participants via a brief questionnaire administered through Research Electronic Data Capture (REDCap) (26, 27).

#### Interview data collection

Semi-structured interviews were conducted with patients between June and December 2023 to explore experiences related to home BP monitoring and hypertension management (23). The interview guide (Supplementary File 2) was developed and iteratively refined by the research team to include questions about motivations for self-monitoring, steps taken to measure BP, education received, information accessed, and preferences for patient education strategies.

Semi-structured interviews with healthcare practitioners were conducted between August 2024 and 2025 as part of a separate study investigating clinical workflows, delivery of patient education, and barriers and facilitators to hypertension management across primary care settings, including contributions to team-based care (interview guide, Supplementary File 3). For the current study, a subset of these interviews was selected to validate the patient personas.

All interviews were conducted once via Zoom or Microsoft Teams, lasted up to one hour, and were recorded and auto-transcribed verbatim. Interviews were conducted by researchers trained in qualitative methods (KS, EC, GS, RT, FL) and were attended by one-two researchers and the participant. No prior relationship existed between participants and interviewers. The research team met regularly to review data collection processes and field notes, with interview schedules refined iteratively to explore emerging themes. Interviews continued until theoretical saturation was reached (28) for topics related to hypertension management and patient education. Transcripts were not returned to interview participants.

### Data analysis

Figure 1 illustrates the sequential process used to identify barriers and enablers to patient education for hypertension management across time-points in the hypertension care journey and to synthesise these behavioural patterns into patient personas.

**Figure 1.**
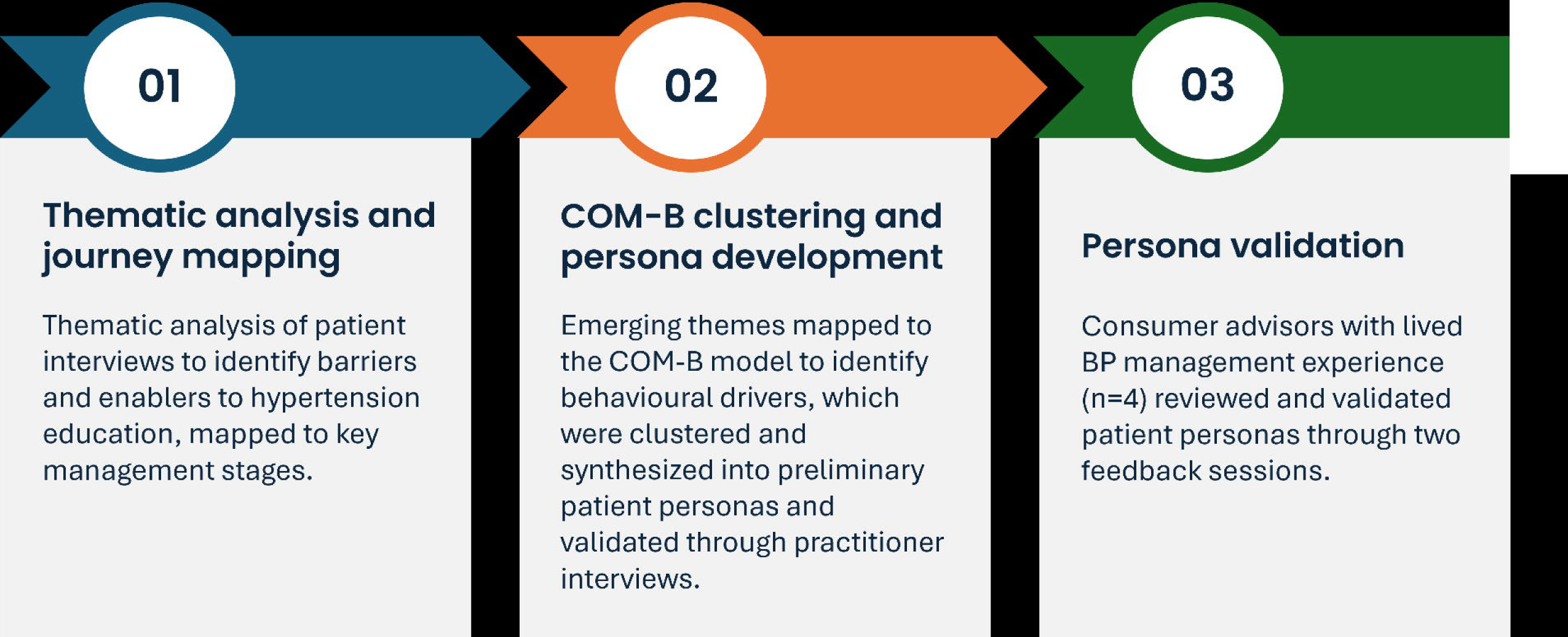
Analytic process for identifying barriers and enablers for patient education and developing patient personas. BP: blood pressure. COM-B: Capability, Opportunity, Motivation-Behaviour Model

### Demographic information

Descriptive statistics were used to analyse categorical demographic characteristics.

### Step 1: Thematic analysis and journey mapping

Semi-structured interviews with patients were analysed using Framework Analysis (29) in Nvivo (V 1.7.1 (1534)). A deductive coding framework was constructed to categorise participant answers according to the interview questions and the components related to patient education for BP measurement and management, such as the topics of education, where education was accessed, who was involved in education delivery and patient perspectives on education (Supplement File 4). One independent study investigator (FL) thematically analysed all interview transcripts using the coding framework. A subset of interview transcripts was coded by two additional study investigators (KS, EC), discrepancies between all coded transcripts were discussed with study investigators (FL, KS, EC) and changes were made to the coding framework where appropriate to incorporate emerging themes not captured in the initial structure.

Following thematic coding, patient journey mapping was used to organise and interpret the data across the hypertension management journey, specifically identifying how patient education needs and behaviours varied at different stages. These stages were organised into diagnosis, treatment initiation and long-term management, which aligned with clinical actions and decision points outlined in international hypertension guidelines (3, 30) to ensure the mapping reflected real-world care pathways. This process identified recurring education-related challenges at each stage, which were then synthesised into broader themes of patient education (such as delivery of education, attitudes to education and health literacy) and hypertension management (such as steps to BP measurement, anti-hypertensive medication, lifestyle modification). Participants did not provide feedback on the findings.

### Step 2: COM-B clustering and persona development

Patient education themes were deductively mapped to the Theoretical Domains Framework and Capability, Opportunity Motivation – Behaviour (COM-B) model by one study investigator (KS) and independently reviewed by another (EC) using a structured mapping matrix developed by the research team (Supplement File 5). The Theoretical Domains Framework identifies specific behavioural determinants across 14 domains (31), while COM-B provides an overarching structure for understanding how capability (an individual’s psychological and physical capacity to engage in the behaviour), opportunity (external factors that enable or prevent the behaviour), and motivation (internal processes that influence decision-making and behavioural direction) interact to influence behaviour (32). Used together, these frameworks offered a comprehensive approach to mapping and enabled the identification of behavioural drivers and barriers underpinning patient education needs at different stages in the patient journey (diagnosis, treatment initiation, and long-term management). These behavioural drivers were then used to inform the clustering of patient experiences, which were synthesised into preliminary patient personas, with each persona developed as a coherent narrative integrating behavioural patterns and relevant contextual factors. While some behaviours (e.g., anxiety at diagnosis, incomplete information provided by health professionals to perform a task) appeared across multiple personas, clustering allowed us to group related experiences into distinct profiles that reflect real-world variability. Demographic characteristics of interview participants including age, gender, ethnicity, health literacy and socioeconomic background were also incorporated into the personas to reflect the diversity of patients represented in the data. Study investigators (KS, EC) met regularly to discuss findings and refine the preliminary patient personas.

To assess the relevance and completeness of the preliminary personas, a subset of semi-structured practitioner interviews (n=12) was independently reviewed by two researchers (KS, EC). Each researcher read through transcripts (n=6 per researcher) and extracted quotes related to the types of patients these practitioners commonly encountered, as well as their experiences with patient education and engagement in hypertension management (Supplementary File 5). These quotes were deductively mapped against the preliminary personas in a structured template. Where practitioner data revealed experiences not fully captured in preliminary personas, such as co-morbidities experienced and BP measurement type, amendments were made to ensure the personas reflected real-world complexities. All amendments were discussed and confirmed by both researchers and further input was sought from investigators who are also practising clinicians (n=3; BK, LKO, CS) to refine the patient personas.

### Step 3: Persona validation

Consumer advisors with lived experience of BP management (n=4) met on two occasions (one hour in duration) to provide feedback on the patient personas. The first meeting (May 2025) aimed to familiarise consumers with study methods and the patient persona concept. One study investigator gave a brief overview of study methods, supported by graphics and introduced personas using short summaries of each persona (e.g. name, point in hypertension journey, diagnoses, patient education received) (KS). Discussion was facilitated among consumers to provide feedback on the concept and content of the drafted personas, the usefulness of personas and areas for improvement using their lived experience. Feedback was summarised and applied to the personas. Ahead of the second meeting, detailed persona summaries were sent to consumer advisors via email for independent review to ensure consumer feedback was appropriately applied. The second meeting (August 2025) aimed to gather structured feedback on the relevance and accuracy of personas. To do this, a study investigator facilitated discussion to determine if the personas aligned with the lived experiences of consumer advisors and if key time-points in hypertension care were captured (KS).

This feedback was incorporated into the final versions, and the final six patient personas were reviewed and approved by all members of the authorship team. Both meetings were recorded, and field notes were taken by study investigators (KS, NC, EC). Patient persona images were created using stock photos available in Microsoft PowerPoint. These images are licensed for use without attribution under Microsoft’s standard user agreement. No generative AI tools were used in the creation of these visuals.

## RESULTS

### Participant characteristics

As shown in Table 1, most patient participants (n=27, 41% women) were aged 50 or older (n=21, 78%) and White (n=25, 93%). Most participants had a diagnosis of hypertension (n=20, 74%) and 36% (n=9) had a previous heart attack or stroke. Health professionals (n=12) were predominantly female (n=11, 92%), had <5 years’ experience in their profession (n=6, 50%), were below the age of 60 (n=12, 100%) and primarily working in areas of most advantaged socioeconomic status (n=5, 42%) or most disadvantaged (n=4, 33%).

**Table 1.** Characteristics of participants: Australian adults (n=27) who measure BP at home and health professionals (n=12) who provide care to people who manage hypertension.

| Demographic characteristics | Patients<br>(n=27) | Health professionals<br>(n=12) |
| --- | --- | --- |
|  | n (%) | n (%) |
| **Gender** |  |  |
| Woman | 11 (40.7) | 11 (91.7) |
| Man | 16 (59.3) | 1 (8.3) |
| **Age** |  |  |
| 18-60 years | 14 (51.9) | 12 (100.0) |
| 60 years or older | 13 (48.1) | 0 (0.0) |
| **Ethnicity** |  | Not collected. |
| Caucasian | 25 (92.6) |  |
| East or South Asian | 1 (3.7) |  |
| Other | 1 (3.7) |  |
| **Employment** |  | Not collected. |
| Employed full-time | 13 (48.2) |  |
| Employed part-time | 5 (18.5) |  |
| Seeking opportunities or unemployed | 2 (7.4) |  |
| Retired | 7 (25.9) |  |
| **Years experience in profession** | Not reported |  |
| <5 years |  | 6 (50.0) |
| 5-9 years |  | 3 (25.0) |
| 10 or more years |  | 3 (25.0) |
| **Education** |  | Not collected. |
| University degree | 12 (44.4) |  |
| Year 12 or equivalent (HSC/Leaving certificate) | 1 (3.7) |  |
| Year 10 or equivalent or less | 6 (22.2) |  |
| Vocational qualification | 8 (29.6) |  |
| **Socio-economic Status\*** |  |  |
| Most disadvantaged, lowest socio-economic status | 7 (25.9) | 4 (33.3) |
| Disadvantaged | 5 (18.5) | 0 (0.0) |
| Average | 4 (14.8) | 1 (8.3) |
| Advantaged | 8 (29.6) | 2 (16.7) |
| Most advantaged, highest socio-economic status | 3 (11.1) | 5 (41.7) |
| **Additional risk factors** |  | Not collected. |
| Diagnosed with high blood pressure | 20 (74.1) |  |
| Previous heart attack, heart disease or stroke | 9 (36.0) |  |
| Chronic kidney disease or impaired kidney function | 4 (16.0) |  |

BP: blood pressure. HSC: high school certificate. \*Postcode was used to classify socio-economic status according to the Index of Relative Socio-Economic Advantage and Disadvantage, which summarizes individual
and household economic and social information to create a place-based indicator of relative advantage and disadvantage (26). Socio-Economic Indexes for Areas \(SEIFA\)

### Patient education for hypertension

Along the hypertension journey (diagnosis, treatment and long-term management), patient education for hypertension management was unstandardised in delivery and content. Education was often opportunistic, primarily verbal, and rarely supplemented with resources. Patients frequently reported that education was insufficient in breadth and detail, prompting them to seek information online, especially at long-term management. The alignment of these barriers and enablers with behavioural mechanisms of the COM-B model is outlined in Table 2.

**Table 2.** Experience of patients and healthcare practitioners in receiving and delivering patient education for hypertension management.

| COM-B Domain | Theme | Barrier or enabler | Illustrative quotes |
| --- | --- | --- | --- |
| **Capability**<br>psychological | Language and clarity | Barrier | *“Explain to me the high point... maybe show me the chart of my blood pressure.”* (Patient, female, 50-59 years old) |
|  |  | Barrier | *“So, 140/90 would be my threshold. Although I have to say, like, I think in America, they're using 130/80, maybe and sometimes it's a bit confusing”* (Nurse, female, 30-39 years old) |
|  |  | Barrier | *“Even still today I get a bit confused, and I get varied responses, and it depends on whom I'm talking to now.”* (Patient, male, 40-49 years old) |
|  |  | Enabler | *“She gave me a chart... green, yellow, different levels... if I'm in the normal range I don't need alarm.”* (Patient, female, 40-49 years old) |
|  | Knowledge and prior experience | Enabler | *“I can figure stuff out, whereas a lot of people couldn't figure stuff out and I spent probably 10 to 20 hours on the Internet reading about home blood pressure.”* (Patient, male, 60-69 years old) |
|  |  | Barrier | *“But other things like I think there was eating and drinking and things like that I probably didn't consider how that might influence blood pressure. So that might have been good to know.”* (Patient, female, 20-29 years old) |
| **Capability**<br>physical | Language and health literacy | Barrier | *“He just used all the medical jargon... that language didn't mean anything to me.”* (Patient, female, 60-69 years old) |
|  |  | Barrier | *“If they are culturally and linguistically diverse... it's very hard to get things.”* (GP, female, 30-39 years old) |
|  |  | Enabler | *“You're not going to achieve patient change unless they have health literacy... patient has a better experience when educated.”* (GP, female, 30-39 years old) |
| **Opportunity**<br>physical | Access to care and resources | Barrier | *“I can't afford to go to the doctor all the time... they charge you for Medicare rebate.”* (Patient, female, 60-69 years old) |
|  |  | Barrier | *“If we were trying to find like sort of a leaflet of some sort to give to the patient, then I guess we'd probably just Google something.”* (Pharmacist, female, 40-49 years old) |
|  |  | Enabler | *“The only thing... was having a look on the Internet... what the good blood pressure results were.”* (Patient, female, 60-69 years old) |
|  |  | Barrier | *“Time is a factor for everything that they treat, I think that 10-minute slot.”* (Patient, male, 50-59 years old) |
| **Opportunity**<br>social | Relationships and social support | Barrier | *“He [GP] takes it with a grain of salt, I think. He sort of takes his own measurement and that's what he goes by.”* (Patient, male, 50-59 years old) |
|  |  | Barrier | *“Oh, It's tricky. I don't think they really notify us whether they took [blood pressure] at the pharmacy. I* *haven't encountered any instance of such.* " (Nurse, female, 18-30 years old) |
|  |  | Enabler | *"I learned to do BP because of my father-in-law..."* (Patient, female, 50-59 years old) |
|  |  | Enabler | *"It does make a lot of a lot of difference about a doctor is and how they treat you...and the nurses."* (Patient, female, 50-59 years old) |
| **Motivation**<br>reflective | Perceived risk and patient engagement | Barrier | *"If you don't know why you're doing it, you're less likely to do it."* (Patient, male, 60-65 years old) |
|  |  | Barrier | *"GPs... don't take blood pressure as a serious thing."* (Patient, male, 50-59 years old) |
|  |  | Barrier | *"I really saw that like people that ... multiple comorbidities would just jump between doctors and like none of it was being managed properly."* (GP, female, 30-39 years old) |
|  |  | Enabler | *"I've had to physically tell the health professionals... look, it's high."* (Patient, male, 50-59 years old) |
| **Motivation**<br>automatic | Prompts to behaviour change | Enabler | *"If there's something on TV...where the doctor is measuring blood pressure I'll think, I should really do that."* (Patient, female, 60-69 years old) |
|  |  | Barrier | *"They asked you to measure it and you ring for advice, and they say don't worry about it."* (Patient, female, 60-69 years old) |

Capability (psychological): Widespread confusion and ambiguity around key pieces of information and prior knowledge impacted learning experience. Capability (physical): Health literacy and language impacted perceived capacity to learn. Opportunity (physical): Cost and time are barriers to accessing information from a health practitioner, while online information is accessible. Opportunity (social): Limited communication between healthcare practitioners builds confusion and inefficiency. Motivation (reflective): Misalignment of motivation to manage hypertension between health practitioner and patient creates challenges. Motivation (automatic): support from health practitioners reinforces behaviour.

Barriers and enablers to patient education also varied across the hypertension journey as demonstrated in Figure 2. At diagnosis, patients often faced confusion and uncertainty as they tried to make sense of their condition. Capability was constrained by the use of complex medical jargon and practitioner knowledge gaps, while opportunity was limited by short appointment times which inhibited quality and comprehensive education and clinical inertia around BP thresholds, preventing timely intervention.

> *“He just used all the medical jargon that was incorporated into the report, which you know, I can understand they’re trying to be as accurate as possible, but that that language didn’t mean anything to me. That was terrible. I’ve just paid, you know, however much for a telehealth consult that’s just been …. You could have just sent me the report.”* (Patient, female, 60-69 years old).

**Figure 2.**
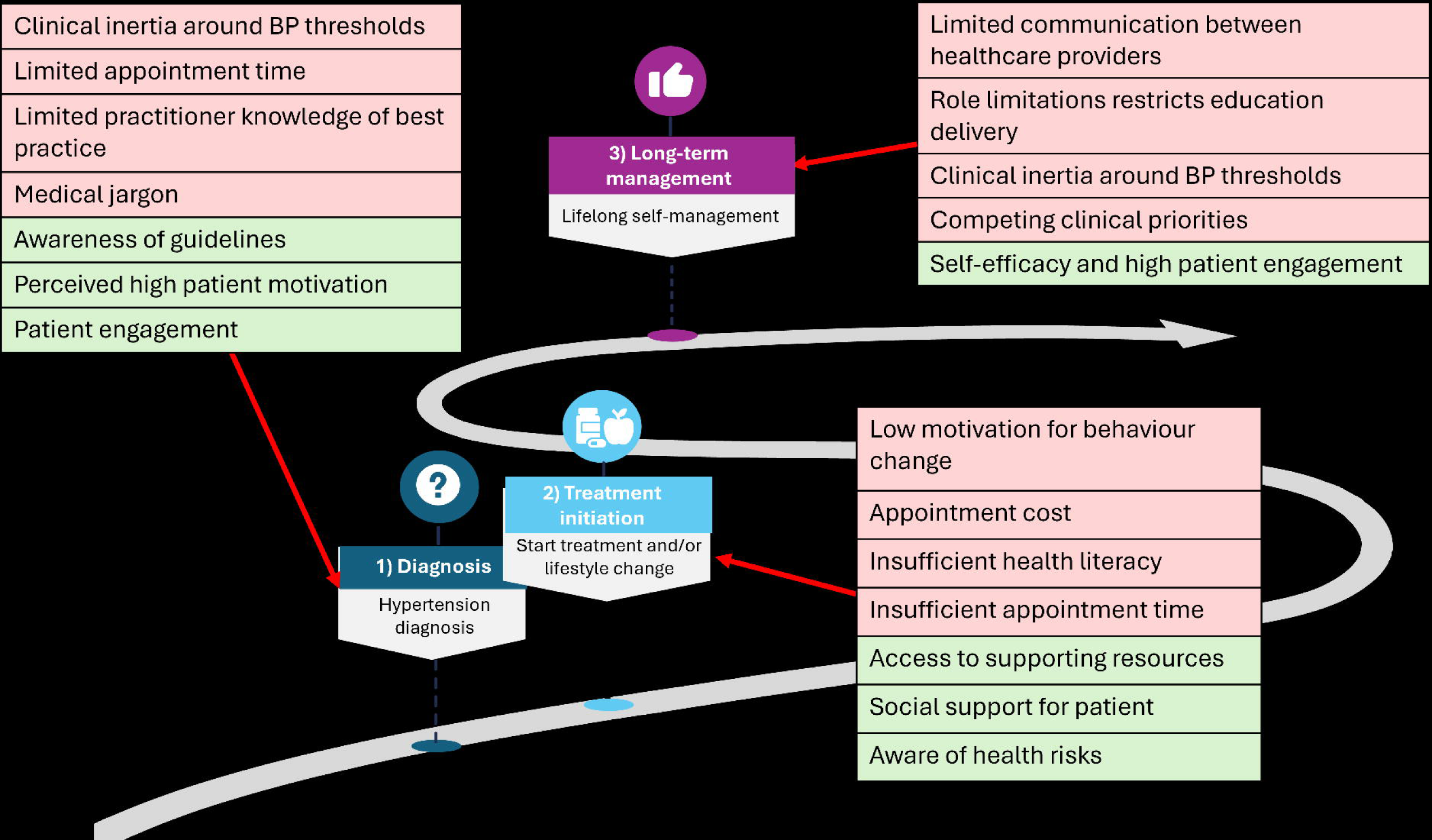
Barriers and enablers to patient education experienced along the care journey. GP. General practitioner. The red shaded boxes represent barriers to patient education, whereas the green shaded boxes represent enablers.

> *“Time is a factor for everything that they treat, I think that 10 minute slot is, you know, it’s a whole Medicare bulk billing. That’s a big whole separate issue.”* (Patient, female, 50-59 years old).

Despite these challenges, high patient motivation and practitioner awareness of guideline recommendations supported early engagement in care.

During treatment initiation, patients navigated the practicalities of starting medication and lifestyle changes, often with mixed clarity and confidence. Capability was reduced by inconsistent messaging, limited health literacy and uncertainty around BP measurement technique and reading interpretation.

> *“And also I thought for a lot of people, including me, if you don’t know why you’re doing it, you’re less likely to do it. I think a lot of the reason that people don’t do what they’re told to do is because they don’t know why they’re supposed to do it so they just don’t, including me.”* (Patient, male, 60-69 years old).

While opportunity was restricted by cost of care and lack of structured education delivery and resources, social support acted as an enabler.

> *“I actually learned to do blood pressure because of my father-in-law. He was sick near the end of his life and he had diabetes and a few other issues, so when I went to bringing his breakfast and his lunch, I’d always cooking breakfast and lunch and I’d always measure his blood pressure while I was there to make sure that, you know, he was OK.”* (Patient, female, 60-69 years old).

Motivation was often low for behaviour change due to competing priorities and the asymptomatic nature of hypertension, though awareness of long-term health risks associated with hypertension encouraged uptake.

In the long-term management phase (typically beyond 12 months after diagnosis), patients worked to sustain routines and interpret ongoing information, often amid fragmented care. Opportunity was impacted by fragmented interprofessional communication which hindered task sharing and competing clinical priorities, although was enhanced through access to bulk-billed services and support from family or community members. Patient motivation and self-efficacy were key drivers of sustained engagement in education and care.

> *“Even still today I get a bit confused, and I get varied responses, and it depends on whom I’m talking to now. I still really don’t know what a good normal reading is.”* (Patient, male, 40-49 years old).

### Patient personas reflecting patient experiences

Six patient personas were developed to synthesise clusters of patient experiences, capturing behaviours and the associated barriers, enablers, and COM-B factors influencing engagement with hypertension education across the care journey. Figure 3 demonstrates an example visual summary of each persona, and summarizes their key management barriers and enablers, and behavioural factors (COM-B) along the hypertension care journey. Detailed summaries of their key characteristics and behavioural barriers and enablers, as well as a summary of each individual persona are included in Supplementary Files 6 and 7.

**Figure 3.**
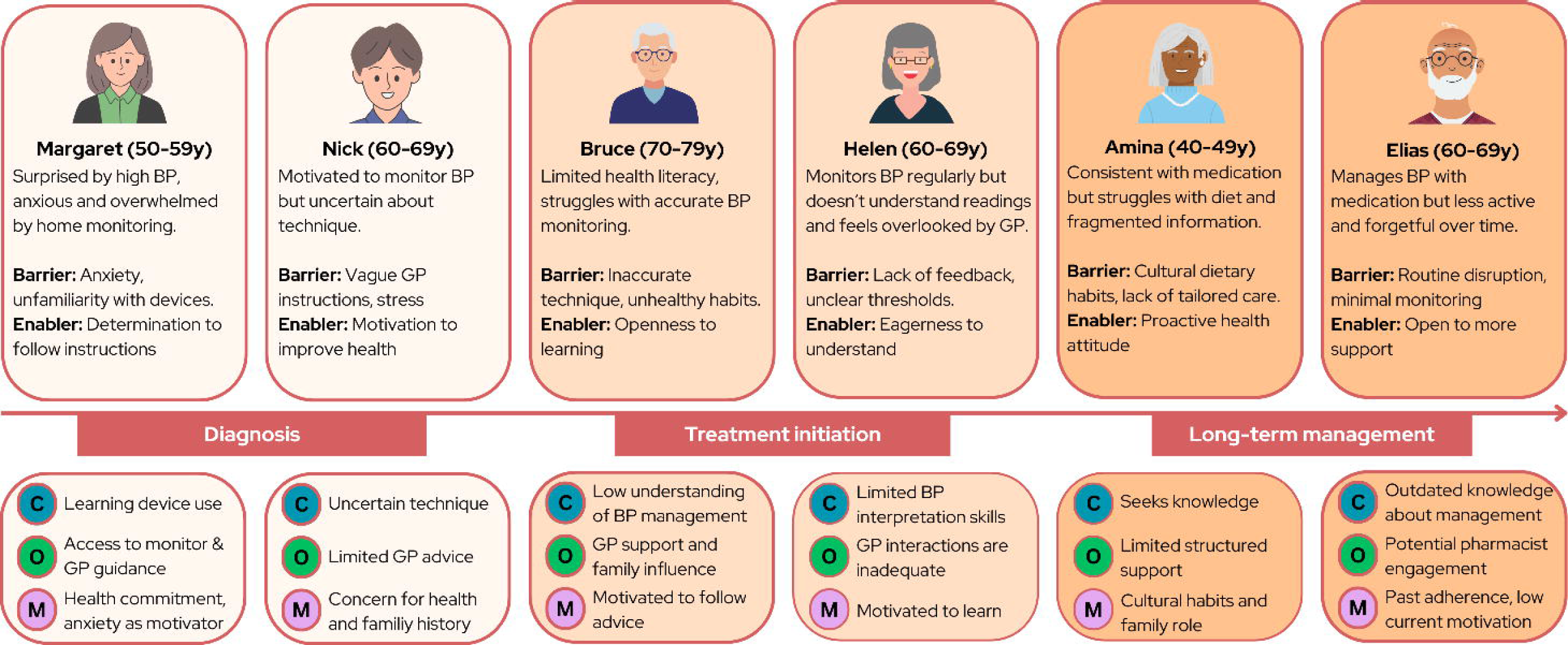
Summary patient personas reflecting the experiences of patients obtaining education for hypertension throughout the hypertension care journey. BP: blood pressure. GP: general practitioner. C: capability, O: opportunity, M: motivation

Across the six personas, recurring challenges were related to common hypertension management behaviours such as medication adherence, lifestyle change, home BP measurement and navigation of multiple health services. Barriers commonly related to limited capability (e.g., uncertainty about home BP measurement technique, low health literacy), constrained opportunity (e.g., fragmented care, lack of structured support), and fluctuating motivation (e.g., competing priorities, assumptions that medication alone was sufficient). Enablers included strong health goals, proactive information-seeking, and supportive relationships.

## DISCUSSION AND CONCLUSION

### Discussion

This study set out to identify key barriers and enablers to patient education across the hypertension journey, from diagnosis through to long-term management. By analysing patients’ experiences at each stage, we identified clusters of behavioural and contextual factors that shape how individuals understand, engage with, and act on hypertension education. These common behavioural patterns were synthesised into six patient personas, which serve to exemplify and translate these findings into a practical, person-centred tool for health professionals and intervention designers. The diversity in the patient experience underscores the need for tailored person-centred strategies that respond to the evolving needs, capacities, and contexts of patients navigating hypertension management in primary care.

Effective hypertension management requires holistic intervention because it is shaped by not only clinical factors but also by emotional, behavioural, and contextual influences (33, 34). Further, it requires patients to have the capability (knowledge, skills, and confidence to measure BP accurately), the opportunity (access to medications, supportive environments, and healthcare services), and the motivation (belief in the value of treatment, emotional readiness) to engage in sustained self-management behaviours (4, 35, 36). The personas developed in this study reflect the multifaceted nature of hypertension across the care journey, capturing how patients navigate uncertainty and fluctuating motivation alongside practical challenges such as interpreting measurement data or integrating lifestyle changes. Emotional responses, such as feeling overwhelmed at diagnosis or frustrated by inconsistent advice often intersect with contextual factors like the cost of healthcare visits, family responsibilities, and cultural expectations, influencing engagement with education and care (5). This complexity is further compounded by the asymptomatic nature of hypertension, which can delay diagnosis and reduce the perceived urgency for treatment initiation and adherence (33).

Patients with hypertension often juggle multiple comorbidities making it difficult to prioritise hypertension management, especially when symptoms are absent or minimal (33). These overlapping challenges also make it difficult for practitioners to accurately identify and address individual patient needs. This underscores the value of patient personas as a tool to visualize and communicate diverse patient experiences and behavioural drivers to inform interventions that are not only clinically effective but also contextually appropriate and practically feasible. Beyond education, personas also have clinical utility in health provider training and care planning, enabling practitioners to better engage with patients lived experiences and tailor care accordingly.

Hypertension self-management can be challenging for patients due to limited health literacy and inconsistent support across care settings (15, 37, 38). Health literacy has been shown to influence not only the ability to understand and act on health information, but also motivation, self-efficacy, and problem-solving skills, all of which are central components to effective hypertension management (38). Patient health literacy levels also directly impact one’s learning needs, such as complexity of information and language, use of supporting written materials and follow up (5). Many participants in our study described receiving education that was inconsistent between providers, overly technical, or failed to meet their individual needs, demonstrating that when education is not tailored to individual needs, it can create barriers to engaging in care. This aligns to previous work demonstrating that healthcare providers are not trained to identify and adapt for patient health literacy needs (4, 39) and patient education that relies on medical jargon or assumes a one-size-fits-all approach fails to empower patients (40). Therefore, the personas developed in this study offer a practical tool to bridge this gap, highlighting how health literacy manifests in real-world hypertension management and associated patient needs and preferences for education. They highlight the diversity of patient experiences and inform education strategies that are both behaviourally informed and responsive to patients lived realities and needs.

Importantly, the personas highlight opportunities to strengthen team-based approaches to patient education by identifying critical time points in the hypertension journey where coordinated, tailored education is most needed and can be improved. This includes at diagnosis, when patients are often confused and anxious, during treatment initiation, when decisions about medication and lifestyle changes are made, and in long-term management, where sustaining motivation and adherence becomes a central challenge. These insights highlight the need for education strategies that are not only timely and relevant but also responsive to the lived realities of patients across the hypertension care journey. Primary care is central in hypertension management, however barriers within this setting often hinder effective patient education, such as limited patient and provider motivation, cultural and linguistic differences, time and funding constraints and limited communication and coordination between settings (33, 41, 42). The personas revealed how fragmented communication between GPs, nurses, and pharmacists can lead to inconsistent messaging, or missed opportunities for timely intervention. Time constraints in consultations, assumptions about patient motivation or literacy, and limited follow-up mechanisms further compound these challenges. These insights point to the need for a more structured, team-based approach to patient education, with clearly defined roles across different stages of the hypertension journey and targeted learning outcomes to guide delivery (43, 44). Therefore, with the growing emphasis on patient-centred care, it is critical that the delivery of patient education in primary care is consistent, coordinated, and grounded in lived experiences to meaningfully support engagement and improve health outcomes.

#### Strengths and limitations

This study has several limitations that should be acknowledged. First, while the sample included 27 patient interviews and 12 practitioner interviews, all participants were based in Australia, which may limit the generalisability of findings. However, international guidelines for hypertension management share core principles and, although minimally varied in structure, aim to achieve similar outcomes (3, 30, 45). Therefore, while contextual differences exist, the patient hypertension journey tends to follow broadly similar pathways globally and the behavioural insights and barriers identified in this study are likely to resonate with patients in other settings. The demographic scope of participants, however, lacked strong representation from culturally and linguistically diverse communities, which may have excluded important perspectives on navigating hypertension care. Second, the interview guide was designed primarily around experiences of uncomplicated hypertension and primary prevention. As such, the personas may not fully capture the complexities faced by patients with multiple comorbidities or those managing polypharmacy, which are common in real-world hypertension management. To mitigate this and enhance the validity and relevance of the personas, we triangulated patient data with insights from practitioner interviews, and consultation with clinician-researchers working in primary care and consumer advisors. This multi-perspective approach strengthens the validity of the personas, though further testing in more diverse populations and settings is warranted. Further, by using framework analysis to structure persona development, this study not only captures the lived experience of hypertension management but also lays the groundwork for designing targeted, theory-informed interventions that are responsive to behavioural needs (32, 46).

### Practice implications

This study offers a structured approach to understanding the diverse needs, motivations, and barriers patients face when managing hypertension to guide the delivery of tailored education strategies in primary care. Beyond education, personas have clinical utility in training, care planning, and enhancing communication strategies, supporting practitioners to better engage with patients lived experiences. Future directions include testing the integration of these personas into primary care intervention design, clinical workflows, and digital tools, as well as adapting them for other chronic conditions.

## Conclusions

Findings from this study provide insight into the behavioural, emotional, and contextual realities of hypertension management in primary care through the development of evidence-based patient personas. By embedding the COM-B model into persona development, we captured the complexity of hypertension self-management in a way that is both theoretically grounded and practically useful.

## Data availability

The data that support the findings of this study are available from the corresponding author upon reasonable request.

## Author contributions

Conceptualisation, KS, EC and NC.; methodology, KS, EC and NC; formal analysis, KS, EC and FL; writing – original draft preparation, KS, EC; writing – review and editing, KS, EC, CB, EH, BK, LKO, LL, FL, MS, GS, MS, CS, RT, AES and NC; supervision, NC; project administration, KS. All authors have read and agreed to the final version of the manuscript.

## Acknowledgements.

We would like to sincerely thank our consumer advisors, John Stevens, Kim Beesley and William Steele, as well as one advisor who preferred to remain anonymous for their invaluable insights, feedback, and contributions to this work. Their perspectives greatly informed the development, refinement and applicability of the implementation strategies.

## SUPPLEMENTARY MATERIAL

**Supplementary File 1.** COREQ Checklist

**Supplementary File 2.** Patient interview schedule

**Supplementary File 3.** Practitioner interview schedule

**Supplementary File 4.** Coding Framework

**Supplementary File 5.** COM-B Patient Mapping

**Supplementary File 6.** Summary Patient Personas and COM-B behavioural barriers and enablers

**Supplementary File 7.** Detailed Individual Patient Personas

**Supplementary File 8.** Plain language summary

